# Assessing the relative contributions of healthcare protocols for epidemic control: an example with network transmission model for COVID-19

**DOI:** 10.1101/2020.07.20.20158576

**Authors:** Matheus Tenório Baumgartner, Fernando Miranda Lansac-Tôha

## Abstract

The increasing number of COVID-19 cases threatens human life and requires retainment actions that control the spread of the virus in the absence of effective medical therapy or a reliable vaccine. There is a general consensus that the most efficient health protocol in the actual state is to disrupt the infection chain through social distancing, although economic interests stand against closing non-essential activities and poses a debatable tradeoff. In this study, we used an individual-based age-structured network model to assess the effective roles of different healthcare protocols such as the use of personal protection equipment and social distancing at neighbor- and city-level scales. Using as much as empirical data available in the literature, we calibrated a city model and simulated low, medium, and high parameters representing these protocols. Our results revealed that the model was more sensitive to changes in the parameter representing the rate of contact among people from different neighborhoods, which defends the social distancing at the city-level as the most effective protocol for the control of the disease outbreak. Another important identified parameter represented the use of individual equipment such as masks, face shields, and hand sanitizers like alcohol-based solutions and antiseptic products. Interestingly, our simulations suggest that some periodical activities such as going to the supermarket, gas station, and pharmacy would have little contribution to the SARS-CoV-2 spread once performed within the same neighborhood. As we can see nowadays, there is an inevitable context-dependency and economic pressure on the level of social distancing recommendations, and we reinforce that every decision must be a welfare-oriented science-based decision.

## Introduction

Epidemics usually pose challenges to society by threatening human life, which frequently leads to social disruption and economic depletion (Meltzer et al., 1999). The most recent coronavirus disease (COVID-19) outbreak, caused by the severe acute respiratory syndrome [SARS]-CoV-2 virus, is a particularly urgent global event that already induced massive losses to human life and affected the economic development worldwide (Anderson et al., 2020; Baldwin and Mauro, 2020; Kabir et al., 2020). Since the first case of COVID-19 in Wuhan, Hubei province of China, the disease has established local transmissions in many countries, with the number of confirmed and fatal cases growing exponentially in several regions (Chinazzi et al., 2020; Wilder-Smith et al., 2020).

The rapid spread of this new coronavirus has motivated numerous studies on its epidemiological characteristics (Adhikari et al., 2020; Lipsitch et al., 2020; Rothan and Byrareddy, 2020). Clinical symptoms include high fever, dry cough, and respiratory distress (Lai et al., 2020). However, the disease onset may occasionally turn into severely progressive lung failure owing to alveolar damage (Xu et al., 2020). Clinical characteristics and the common course of ill patients carrying COVID-19 include absent to mild symptoms and are certainly context-dependent, but there is a considerable proportion of individuals that likely require medical assistance, especially those elderly people and/or with underlying comorbidities (K. Liu et al., 2020; Yang et al., 2020). In fact, a common concern for health systems is that an uncontrolled outbreak is definitely catastrophic, and really effective retainment measures are now the only realistic option to avoid the total collapse of healthcare facilities (Adams and Walls, 2020).

There is an ongoing debate about the optimal mitigation strategies to prevent or reduce contagion among people. Strategies range from the use of physical barriers (e.g., masks), hand hygiene, avoidance of direct contact among people (e.g., handshakes and hugs) and social distancing at different scales such as staying meters apart from each other and traveling restrictions (Hellewell et al., 2020; Leung et al., 2020). However, the proposal of distancing interventions on a large scale, such as the suspension of classes and the lockdown of non-essential activities such as many commercial facilities, has been proved as a drawback to the economic interests (Ayittey et al., 2020; Bonaccorsi et al., 2020). This side effect stands against the social distancing protocols, highlighting that the optimal combination of retainment strategies needs to be a welfare-oriented consensus between healthcare and economic sustainability (Gostin and Wiley, 2020). In this context, modeling the epidemic propagation focusing on these strategies can provide useful insights to guide field interventions and to understand COVID-19 infection states (Ferguson et al., 2020). Qualitative information from these models can assist decision-makers and support critical intervention policies.

It is a common perception that the novel coronavirus is transmitted under a contact network among humans (Guo et al., 2020; Rothan and Byrareddy, 2020). The spread of the virus in human networks occurs over time and across the geographical space. Therefore, enhanced models should account for such spatiotemporal dynamics, as well as the individual-level epidemiological phenomena (Li et al., 2019). Moreover, because susceptibility and mortality rates from COVID-19 are age-dependent (Li et al., 2020; Moghadas et al., 2020), nearly optimal epidemiological models must attempt to incorporate these heterogeneities in order to produce more realistic results (Bian, 2004).

Those semi-mechanistic models that define population dynamics considering age groups are known as individual-based age-structured network models (Ajelli et al., 2010). In this endeavor, we built city-level simulations to investigate which strategy could maximize the mitigation of the infection outbreak. Models were calibrated with an empirical spatial structure and specific parameters of COVID-19, considering an extended susceptible-infected-recovered (SIR) epidemiological structure. In depth, we aimed to investigate the relative roles of health protocols such as the direct exposure to SARS-CoV-2, as well as the social distancing on both local and large scales. By varying model parameters related to these protocols, we were able to discuss better scenarios considering the delay in the infection peak and lower numbers of cases, as well as activities with a low potential to boost the outbreak. Our simulations indicate that changes in a single public protocol (e.g., social distancing and individual-level care) could result in quite different patterns of the infection wave. Meanwhile, we reveal that the carrying capacity of healthcare facilities will likely be overloaded and that social distancing, allied with investments in mass testing and hospital facilities, are the most appropriate engagements against COVID-19.

## Results

We calibrated the simulations using an epidemiological model that considered all combinations of relatively low, intermediate, and high probabilities of personal exposure to SARS-CoV-2 (*β*), as well as the probability of contact among people in local (*v*), and regional (*k*) scales. As long as *β, v*, and *k* increases, the individual-level chances of being exposed to the virus and the encountering probability on local and regional scales also increased, respectively.

In this sense, it was clear that there was a trend of faster and higher infection peaks as long as the values of the three parameters increased from scenarios S1 to S27 (Figure 1; Table S1). For those models parametrized with high values (i.e., models with two or three red dots in Figure 1), the infection outbreak peaked 6-8 weeks after the first case, on average. While these scenarios yielded infection waves with many infected individuals already in the first weeks, their peaks were the narrowest though (Table S1). Given the specified model structure, those results forecasting early wave peaks emerged under moderate to high probabilities of the individual-level exposure to SARS-CoV-2 virus (high *β*), in combination with higher encountering rates among people (*v* and *k*) (Figure 1; Table S1).

**Figure 1.**
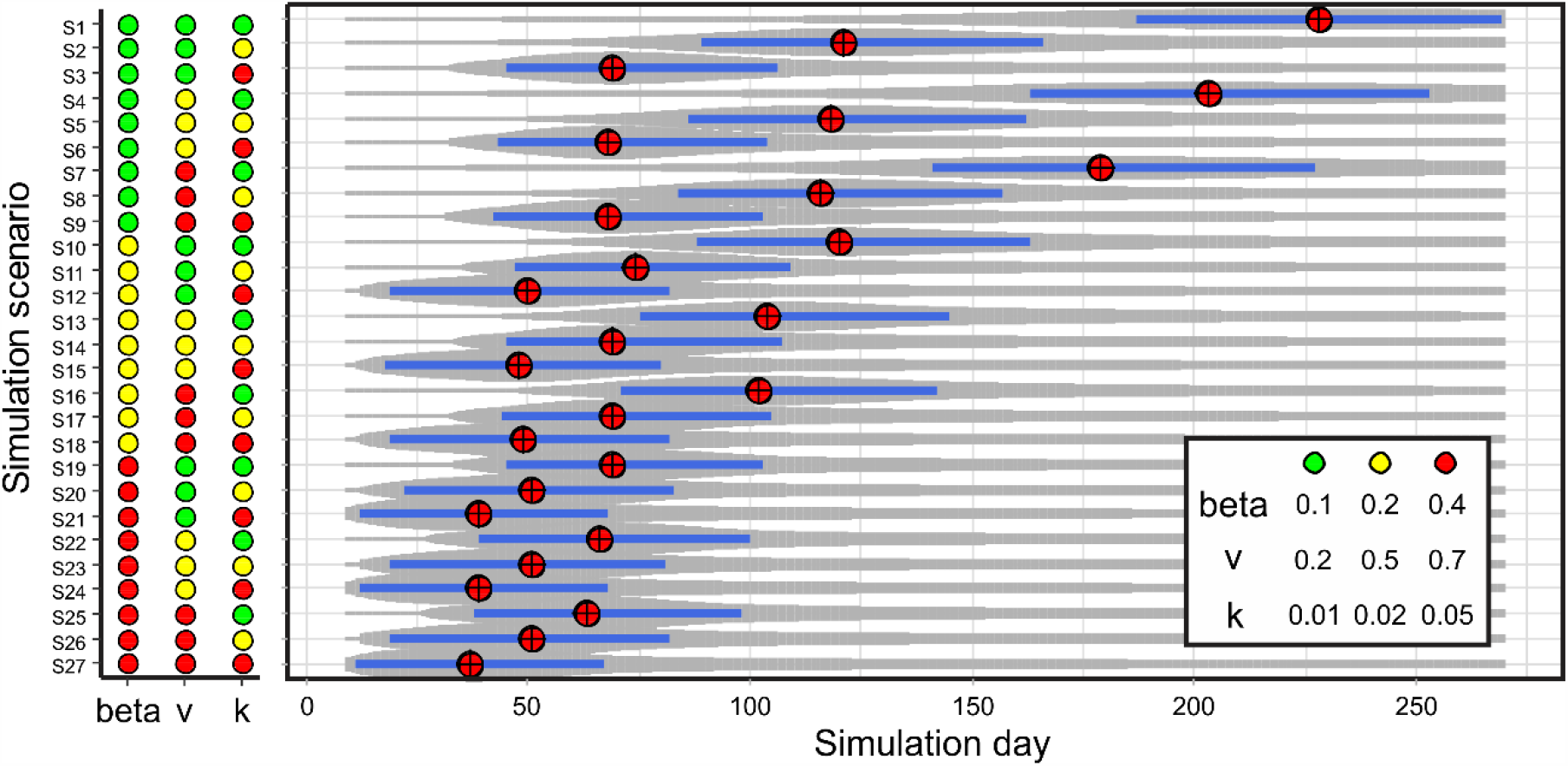
Outcomes of the simulations of COVID-19 infection curves. On the left: green, yellow, and red circles indicate low, intermediate, and high levels of model parameters *β, v*, and *k* for each of the 27 simulation scenarios. Parameter values are depicted in the box on the right panel. On the right: grey shades portray the progression of the infection across simulation time steps, which was obtained through the smoothing of all simulations under each scenario. The width of each shade is scaled to the height of each infection wave. Blue horizontal bars represent the period when the healthcare system is simulated as overloaded, according to empirical data on available beds. Red circles with crosses portray the peaks of infections.

Infection waves peaked later when models were calibrated with lower values of the three parameters. Under these circumstances, waves peaked nearly two times (14-16 weeks) after the first case, on average, when compared to the aforementioned scenarios (Figure 1; Table S1). These scenarios produced flattened infection curves and later peaks, especially when the model assumed that people were less exposed to the virus (low *β*) and had a low probability of encountering at both local (low *v*) and regional (low *k*) scales. Considering our model city, all scenarios potentially overloaded the nominal carrying capacity of the healthcare system (Figure 1). Nevertheless, there was a clear trend that those simulations with steeper smooth waves (bottom scenarios in Figure 1) had a short-lasting overwhelm of the modeled hospital capacity.

To accurately identify the relative effectiveness of each of the mitigation strategies against COVID-19, we modeled the outputs of simulations as a function of different levels of parameters. When we considered the results in terms of the rapid growth of the infection (i.e., day of infection peak, the ratio of total population infected by the virus, and the number of infected people), there were significant influences when increasing parameters *β* and *k* (Table 1). In depth, these results point towards consistent effectiveness against the disease burden by decreasing both the exposure rate of individuals and increasing social distancing on a city-level scale (Table 1) and that this relationship seems to be non-linear (Figure 2). We found that those models calibrated with low exposure rate and high social distancing on a large scale had delayed infection peaks and less infected people (Table 1; Figure 2).

**Table 1.** Partial regression coefficients obtained from multi-response models through canonical powered partial least squares (CPPLS) regressions. The predictor matrix considered all three model parameters (*β, v*, and *k*). We built separate models for each response matrix: infection peak (peak day, the ratio of infected people and the number of cases at the peak) and healthcare saturation (first and last day, duration of the period, and estimated deficit in the number of hospital beds). Significant values under Jack-knife *t*-tests (considering α = 0.01) are in bold. See *Methods* for details.

| | Parameter Estimate | Std. Error | D.f. | $t$ | $P$ |
| --- | --- | --- | --- | --- | --- |
| **Infection peak** |  |  |  |  |  |
| Day |  |  |  |  |  |
| $\beta$ | -0.624 | 0.135 | 26 | -4.619 | **0.000** |
| $v$ | -0.029 | 0.122 | 26 | -0.241 | 0.811 |
| $k$ | -0.560 | 0.112 | 26 | -4.980 | **0.000** |
| Ratio |  |  |  |  |  |
| $\beta$ | 0.645 | 0.122 | 26 | 5.272 | **0.000** |
| $v$ | 0.030 | 0.115 | 26 | 0.264 | 0.794 |
| $k$ | 0.579 | 0.111 | 26 | 5.193 | **0.000** |
| Num. of cases |  |  |  |  |  |
| $\beta$ | 0.646 | 0.123 | 26 | 5.270 | **0.000** |
| $v$ | 0.031 | 0.115 | 26 | 0.267 | 0.792 |
| $k$ | 0.577 | 0.112 | 26 | 5.164 | **0.000** |
| **Healthcare saturation** |  |  |  |  |  |
| First day |  |  |  |  |  |
| $\beta$ | -0.622 | 0.120 | 26 | -5.202 | **0.000** |
| $v$ | -0.085 | 0.122 | 26 | -0.696 | 0.493 |
| $k$ | -0.577 | 0.123 | 26 | -4.680 | **0.000** |
| Last day |  |  |  |  |  |
| $\beta$ | -0.631 | 0.115 | 26 | -5.500 | **0.000** |
| $v$ | -0.071 | 0.116 | 26 | -0.614 | 0.545 |
| $k$ | -0.578 | 0.120 | 26 | -4.833 | **0.000** |
| Duration (days) |  |  |  |  |  |
| $\beta$ | -0.635 | 0.109 | 26 | -5.846 | **0.000** |
| $v$ | 0.001 | 0.110 | 26 | 0.012 | 0.991 |
| $k$ | -0.550 | 0.113 | 26 | -4.852 | **0.000** |
| Deficit |  |  |  |  |  |
| $\beta$ | 0.643 | 0.116 | 26 | 5.533 | **0.000** |
| $\nu$ | 0.048 | 0.114 | 26 | 0.421 | 0.677 |
| $k$ | 0.579 | 0.121 | 26 | 4.802 | **0.000** |

**Figure 2.**
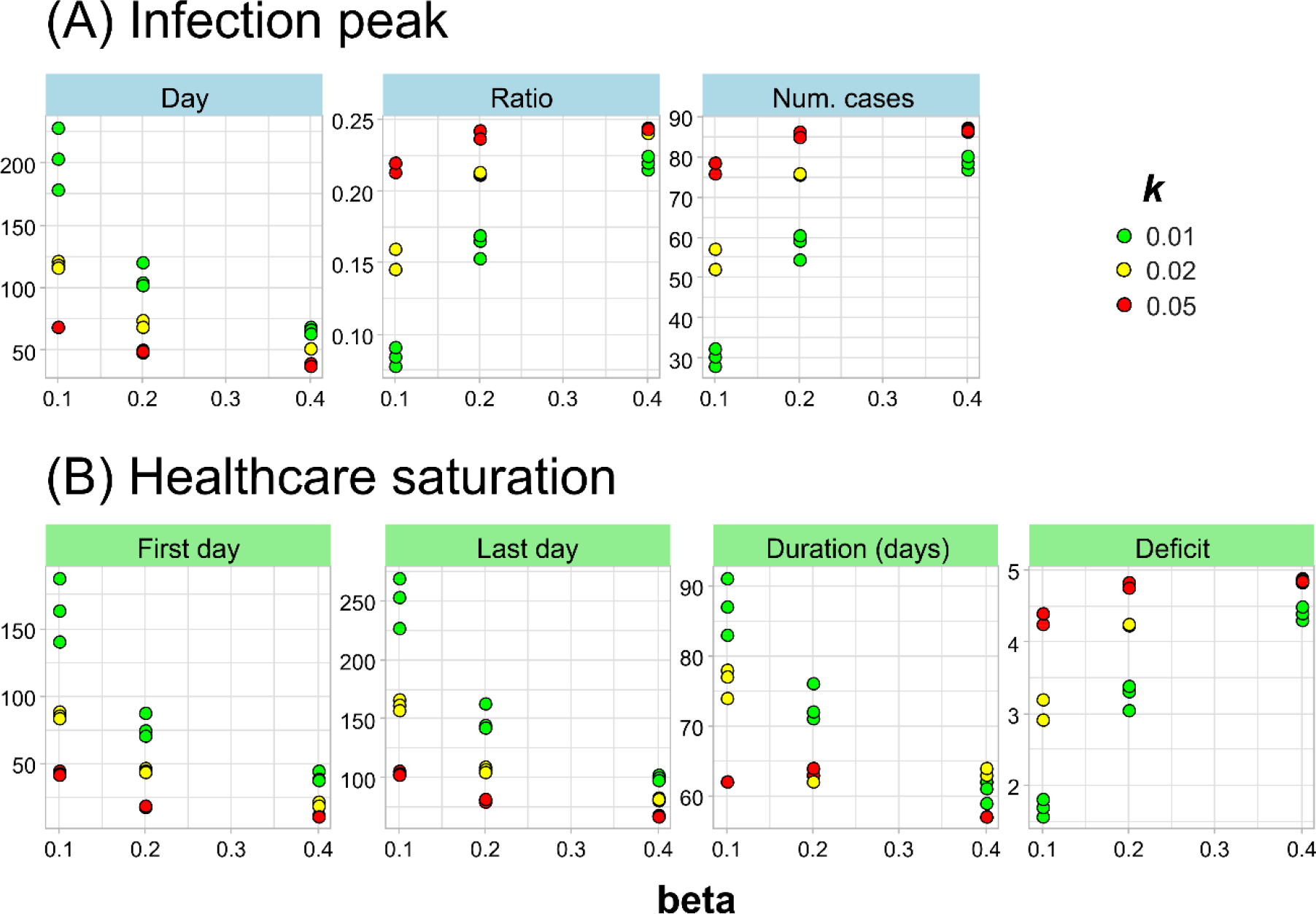
Relationships between model parameters and the response variables. (A) Infection peak; (B) Healthcare saturation.

Similarly, in terms of saturation of the healthcare system (i.e., first, last day and duration of the overwhelm, and the proportional deficit in the number of beds), the model indicated that both exposure rate (*β*) and social distancing at a large scale (*k*; i.e., city-level) also had significant influences (Table 1). All aspects of the infection wave related to the time that the health system would be saturated decreased with both exposure rate and social approximation of people (Figure 2B). These scenarios reveal that the infection waves could be dramatically earlier and intense if people get infected at an increased rate. However, the predictions also reveal more dramatic forecasts, where the number of healthcare units should likely be three to fivefold the number of potentially available beds (Figure 2B). The results reveal no significant effect of the social distancing protocols on a local level (i.e., neighborhood).

## Discussion

As an increasing number of COVID-19 cases is still being identified due to the impressive transmissibility of SARS-CoV-2 (He et al., 2020; Y. Liu et al., 2020), economic consequences became a major concern (Kabir et al., 2020; McKee and Stuckler, 2020). It is therefore fundamental to determine the relative effectiveness of control measures on disease retainment and to inform decisions about an adequate framework for management and mitigation strategies. For this reason, model projections considering different levels of these protocols are maybe the best coalition between researchers and policy-makers during this epidemic (Ferguson et al., 2020; Kucharski et al., 2020).

It is noteworthy that those countries that took slightly delayed actions (i.e., days or a few weeks after first confirmed cases) had rapid spreads of COVID-19, accompanied by high mortality, which imposed extraordinary demands to the public health systems (Legido-Quigley et al., 2020; Sen-Crowe et al., 2020). Studies demonstrate that strategies became much more effective when combined with the detection and isolation of cases (Anderson et al., 2020; Ferguson et al., 2020; Kretzschmar et al., 2020). Unfortunately, many low-income countries may not afford or are too large to conduct mass testing (Mayorga et al., 2020). Thus, the brunt of retaining the spread of COVID-19 lays on social distancing and associated efforts to manage and control the infection progress (Crokidakis, 2020).

Social distancing and the isolation of infected people is a core intervention protocol for many infectious diseases and acts by reducing the potential for onward transmission, especially when ‘herd immunity’ protocols are unfeasible (Anderson et al., 2020; Lewnard and Lo, 2020). Our simulations under an empirically-parametrized city model confirmed that social distancing, especially limiting inter-neighborhood movements, would have the largest impact on containing the evolution of COVID-19 within cities. There are many ways such as stay-at-home recommendations, home-based offices, and online teaching classes, ranging from voluntary to context-dependent mandatory reasons, which can greatly contribute to reducing the spread of the new coronavirus (Gostin and Wiley, 2020). Our results reinforce that these may be the best current strategies. Notwithstanding, health workers must continue their work, as well as supermarkets, public transportation employees, police, firemen, and others still need to have contact outside their households, where transmission chains may still remain (Kretzschmar et al., 2020). Fortunately, we found that the movements of people within their residential neighborhoods had lower effects on the evolution of the epidemic curves. This likely suggests that some periodically necessary activities, such as going to the supermarket, gas station, pharmacy, bank agency, and others, would have little effect overall, once performed within the same neighborhood.

In our projections, respecting social distancing protocols likely delay and reduce the peak of the infection curve, thereby scattering the number of severe cases over a longer period. More importantly, under this delayed peak, healthcare systems are able to increase their carrying capacity by building up mobile cabin hospitals, which can provide better treatments for ill people and partially reduce the mortality rate (Moghadas et al., 2020). Early actions are fundamental and optimal interventions may precede the overwhelm of healthcare carrying capacity (Prem et al., 2020; Shoukat et al., 2020). As a fortunate example, since March 20, 2020, Maringá (the city used as a model) declared partial lockdown, with considerably reduced traffic of people. The main interventions included the closure of educational institutions and non-essential commercial activities, as well as the complete lockdown from 21:00 pm to 05:00 am for a few weeks. These preventive social distancing protocols were determined only two days after the confirmation of the first case, which contributed even more to increase its effectiveness. However, some cities such as large Brazilian capitals are now experiencing dramatic scenarios, even with early stay-at-home recommendations (Crokidakis, 2020; Dana et al., 2020). Although social distancing may reduce the effective spread of the SARS-CoV-2 virus, it can never be reduced to zero and many people tend to underestimate this protocol since its effects may take weeks to appear (Hellewell et al., 2020).

Equally worrisome is the fact that our projections put the direct exposure of each individual in the frontline of the factors pushing the infection progress towards the worst-case scenarios. These results deal directly with the rate at which citizens are exposed to the virus. The use of personal protective equipment (PPE), such as surgical or fabric-made masks, face shields, and hand sanitizers like alcohol-based solutions and antiseptic products has been strongly recommended to the general public (WHO, 2020). However, this strategy creates a debate. On one hand, the willingness of people to be protected whenever performing any outside activity, or for those who work with essential services and are frequently at moderate to high exposure risks (Bourouiba, 2020). On the other hand, the rational use of this equipment, once the global demand has grown nearly as exponential as the outbreak itself (Feng et al., 2020).

So far, the most effective action seems to be imposing and encouraging the rational use of masks and the offer of hygiene items by decree or other legal dispositions. However, although most of these policies have been adopted in several countries (e.g. Japan, UK, Singapore, and Germany), there is not enough evidence for the real effectiveness of wearing masks alone or in combination with washing hands frequently in preventing the contact- or aerosol-based transmission of SARS-CoV-2 (Feng et al., 2020; Y. Liu et al., 2020; Rothan and Byrareddy, 2020). Besides, the incorrect use of PPE is thought to be worse than not using at all, as well as exaggerated acquisition and overpricing of PPE could be similarly adverse. In this sense, we particularly recommend that people should use PPE adequately, especially when there is potential to spread or get in contact with droplets in the air. Nevertheless, we underline that social distancing protocols seem considerably more effective.

Just like any model, ours have limitations as well. First, we used a fixed number of hospital beds, which is certainly unrealistic if we consider that there is a current effort to expand the nominal carrying capacity of these facilities in many cities (Croda et al., 2020). However, no matter what estimate we use to forecast the deficit in the number of available beds, projections show that there will not be enough ventilators to treat COVID-19 patients in the next few months, even in the best-case scenarios (Ranney et al., 2020). Second, we did not account for time-varying or dynamic public health protocols. As we can see nowadays, there is an inevitable context-dependency and economic pressure on the level of social distancing recommendations. Certainly, models with a real-time structure accounting for this dynamic would be more appropriate to build an evidence-based political framework.

## Methods

### Modelling framework

We used an individual-based age-structured network model with an underlying modified susceptible-infected-recovered (SIR) epidemiological structure. The modeling approach starts with two main components, the node transition graph and the contact network (Fig. S1). The node transition graph consists of five compartments: susceptible (*S*), exposed (*E*), infected (*I*), recovered (*R*), and deceased (*D*). Each individual may only be in one of these compartments at each time and the rate of transition from one to the next is modulated by parameters *β* (transmission rate), *δ*(infection rate), *γ* (recovery rate), and *θ* (mortality rate). The sequence of the modeled progression of COVID-19 infection assumes that each individual may transit from susceptible (*S*) to infected (*I*) and then the model estimates when they are able to recover from a given condition.

The contact network is represented by the number of individuals (*N*; i.e., circles/nodes) as well as by their interactions (i.e., edges/links), whenever an opportunity for transmission arises (Fig. S1). Theoretically, we assume that the interaction between two nodes occurs whenever there is potential contact between individuals (e.g., hugs, handshakes, or airdrops), or individuals may interact with previously infected objects (e.g., doorknobs, handrails, and elevator panels). Thus, these opportunities serve as windows for the spread of the virus from an already infected individual to a new potential host.

In practice, we explicitly modeled each individual and its probabilities of movement through the node transmission network. At first, all individuals were assigned as susceptible (*S*), because of no-known previous immunity against COVID-19 (Shi et al., 2020). Thereafter, the transition of each node to the exposed (*E*) compartment depended on the transmission rate *β* and the combined proportion of infected individuals, both nearby and in potential neighborhoods (*Y*_*i*_). Thus, the probability of a susceptible individual to become exposed to the virus took place at rate *βY*_*i*_. This product forces the transmission probability to be directly proportional to the number of network-level infected individuals, which seems quite realistic. Once individuals enter the exposed (*E*) compartment, the transition to the infected (*I*) stage depends on the infection rate *δ*, which, in practice, portrays the average incubation time (i.e., days until the pathogen will replicate enough so individuals become infectious). At this stage, those recently infected individuals add up to *Y*_*i*_, which increases the overall virulence of the network. Infected individuals would then be transferred to the recovered compartment (*R*) or be unfortunate otherwise (i.e., move to the deceased (*D*) compartment), depending on rates *γ* and *θ*, respectively. The aforementioned structure yielded the following time-dependent Poisson process:

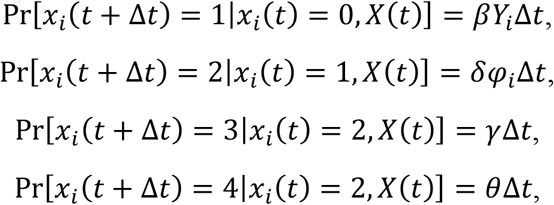

where *x_i_* = {0, 1, …, 4}, which depicts whether the node *i* is susceptible, exposed, infected, recovered, or deceased, respectively (Fig. S1). To include more realism in the model, we included an age-dependent parameter modulating the *E*-to-*I* transition probability (φ_*i*_) that portrays the susceptibility of each *i* individual to become infected at its specific age (Wu et al., 2020; see *Model Parametrization*).

### City structure and movement of people

To incorporate a spatially explicit structure, we modeled the distribution and movement of people based on the city of Maringá, PR, Brazil (23°25’38”S / 51°56’15”W; Fig. S2). This is the seventh largest city in southern Brazil and has a successfully implemented and developed urbanizing plan, which translates into a relatively high efficiency related to the movement of people across the city and alleviates potential traffic bottlenecks. The network city model included empirical demographic information obtained from the 2010 census, consisting of 357,077 citizens (*N*) unevenly distributed across 48 census zones (i.e., neighborhoods; Fig. S2; IBGE, 2010). The city covers approximately 487.7 km^2^ (303 sq mi) in the Northern region of Paraná, South Brazil.

Data for each census zone represented the locations considered in the network model. We extracted the centroid of each zone according to their geographic coordinates (black circles in Fig. S2). To model the movement across the city, we considered that people were assumed to move freely within each location. Under this structure, each individual had an equal probability *v* of contact with all other individuals within the same zone (Sahneh et al., 2017). However, their movement was constrained among locations.

The transmission of the virus from one zone to another was assumed to occur through the movement of individuals across the network. In the city model, for instance, the most frequent reason why people leave their residential neighborhoods is for working purposes (Amram et al., 2019; Wang et al., 2018). Therefore, the spread of the virus resulting from contact among people was proportional to their proximity within the city network, assuming that people tend to work near their own households. This weighting was based on an exponential distance kernel e^-kd^, where *k* scales the probability of individuals from different zones to be in contact, and *d* is the geographic distance (km) between the centroids of each zone.

### Model parametrization

We parameterized the model using as much information as available in the literature. Under our model structure, parameters*δ, γ*, and *θ* were previously approximated and were fixed through all simulations. We then invariably used *δ*=0.196 day^-1^ that portrays 5.1 days of incubation, and *γ* = 0.090 day^-1^, which depicts an 11.1-day period of recovery (Lauer et al., 2020; Pan et al., 2020). The *θ* = 0.00014 value was also fixed, which yielded a fatality ratio of nearly 1.5% (Verity et al., 2020; Wu et al., 2020). We initially calibrated the parameter representing the encountering probability within zones as *v* = 0.7, as suggested in the literature (Sahneh et al., 2017; Sekamatte et al., 2019). Lacking proper knowledge of the remaining parameters, we set *β* = 0.2 and *k* = 0.01. Nevertheless, the model is valid for any value of these parameters (Sekamatte et al., 2019).

### Simulation scenarios

To achieve our objective of comparing different healthcare protocols to retain the spread of the virus, we conducted simulations using different values of the exposure rate (*β*), as well as varying city-level (*k*) and location-level (*v*) probabilities of individuals to get in contact. We then simulated time series based on the aforementioned modified *SIR* model using all possible combinations of parameters *β*{0.1, 0.2, 0.4}, *v* {0.2, 0.5, 0.7}, and *k* {0.01, 0.02, 0.05}. The 27 resulting scenarios considered different combinations of relatively low, medium, and high values of *β, v*, and *k*. This structure was fundamental to isolate the effects of each parameter on the simulated infection waves and to confront the outputs.

We set the model up to start by distributing the 357,077 citizens across the city. To each individual, we assigned a ‘home’ location using the population density of each zone as probabilities (Fig. S2). To include the age-structured information that would approach more realism in simulations, we then randomly assigned an age between 0 and 99 years to each individual, following the empirical age pyramid in Fig. S3 (IBGE, 2010). According to the age of each citizen, we then parametrized the age-dependent infection probability (φ_*i*_) according to Wu et al. (Wu et al., 2020) (Fig. S3). Each simulation started (*t* = 0) with a single COVID-19 case in the Central zone (medical cross in Fig. S2), the location where the first case was recorded on March 18, 2020 in the model city (SMS/MGA, 2020). We then allowed the model to evolve daily until *t* = 270, representing nine months in the future, approximately. For each scenario, we performed 100 simulations. To average the *SIR* curves of all simulations under each scenario, we fitted local polynomial regressions (locally estimated scatterplot smoothing – LOESS) to each node transition compartment, with a smoothing parameter of α = 0.3. From simulated curves, we extracted three response variables related to the infection peak: day of the peak, ratio of infected people, and number of cases at the peak.

### Empirical healthcare data

To infer about prioritizing the efforts of public health personnel, we compared the outcomes of simulations with the nominal carrying capacity of healthcare facilities potentially able to treat COVID-19 cases in Maringá (Fig. S4). We then compared this information with the simulations under all scenarios. We did this because the potential collapse of healthcare systems is a major concern worldwide (Adams and Walls, 2020).

We considered that the number of available hospital beds in Maringá was 1,657, without distinction between ordinary and intensive care units, for simplicity. From these beds, the average normal occupancy is estimated at 58.75% (SMS/MGA, 2020), which yields 684 beds virtually available for COVID-19 cases. Fortunately, most people with infections caused by the SARS-Cov-2 virus only develop mild symptoms and do not require medical care. However, we considered that people would develop severe or critical symptoms at an approximated rate of 0.19 (Wu and McGoogan, 2020). From these severe/critical cases, there is an expected age-dependent probability of hospital admission at 0.025 (0-19 years), 0.32 (20-49 years), 0.32 (50-64 years), and 0.64 (65+ years) (Moghadas et al., 2020; Shoukat et al., 2020). Using these probabilities and the empirical age-structured data from the 2010 census, we extracted the median of a fitted Gamma distribution (Fig. S5) to represent the proportion of infected people demanding hospital beds at each time step (0.0379; interquartile range = 0.0180-0.0697). In the course of each infection wave, we were then able to approximate the duration (i.e., days) of the overload of the carrying capacity of hospitals, as well as a rough deficit in the number of beds available at the projected infection peaks.

### Regression models

Finally, to investigate the relative role of each public health protocol against the spread of SARS-CoV-2 virus, we used multi-response partial least squares regression models (PLS). Specifically, we used the canonical powered extension of PLS (CPPLS), which is suitable for multivariate responses and relationships considering discrete and continuous variables with potential correlation (Indahl et al., 2009). We built models using the varying model parameters (*β, v*, and *k*) as the predictor matrix. As response matrices, we used the three variables related to the infection peak (i.e. peak day, ratio of infected people and number of cases at the peak) and the four variables related to the saturation of the healthcare system (i.e. first and last day, duration and deficit), separately. Variables were centered (i.e., scaled) prior to model fitting to allow for comparisons among parameter estimates. Models were fitted using package ‘pls’ (Mevik et al., 2019) in the R Environment (R Core Team, 2019). The significance of parameter estimates was achieved using approximated Jack-knife *t*-tests (Martens and Martens, 2000).

## Data Availability

All data are included in the manuscript

## Acknowledgements

We would like to thank Marco Túlio Pacheco Coelho, Ricardo Dobrovolski, and José Alexandre Felizola Diniz-Filho for fundamental discussions and suggestions on an early draft of this manuscript. We are also grateful for the infrastructure provided by the Galileo Cloud Computing Program for computational simulations. This work was developed as a scientific counterpart that summarizes many data collected by those people involved directly or indirectly in collecting epidemiologic data and working in favor of health. They deserve all the merit of this paper.

---

**Figure S1.**
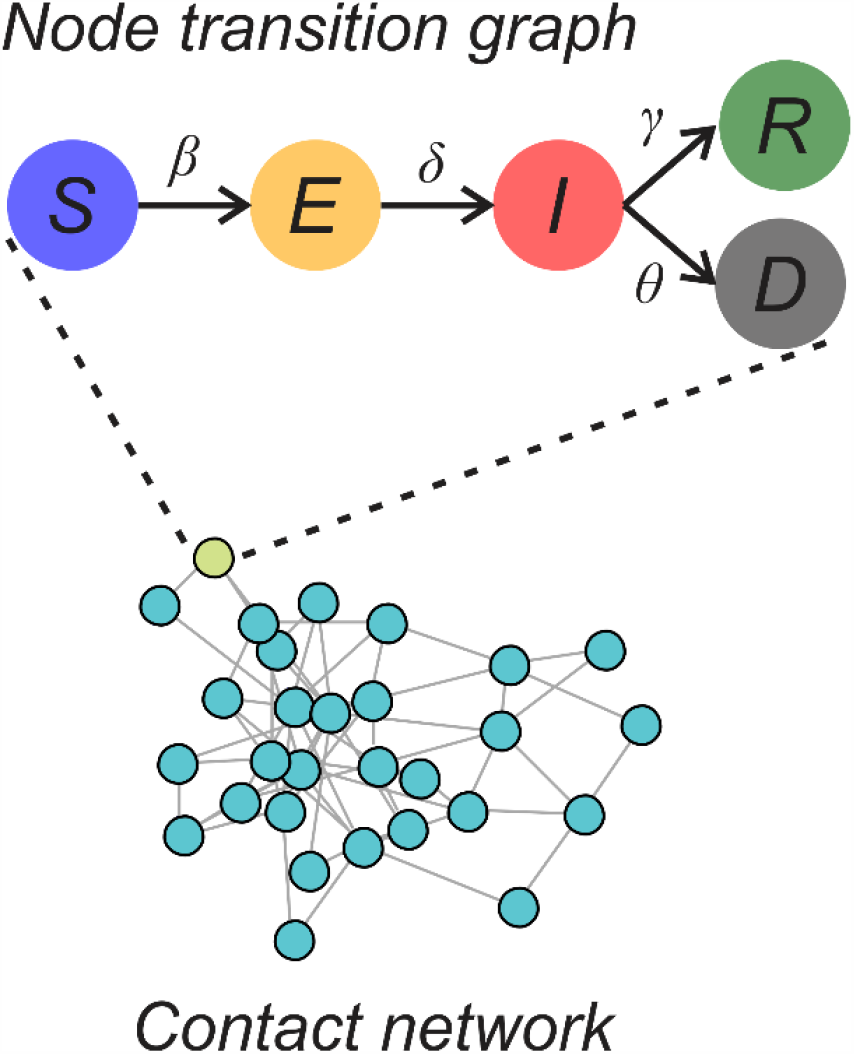
Diagram of the individual-based network model consisting of the transition graph and the contact network. Colored circles represent the five compartments of the model for each node (i.e., individual): susceptible (*S*), exposed (*E*), infected (*I*), recovered (*R*), and deceased (*D*). The transition between compartments is modulated by transmission rate (*β*), infection rate (*δ*), recovery rate (*γ*), and mortality rate (*θ*). Each circle in the contact network represents an individual and links are potential opportunities (i.e., contacts) for COVID-19 transmission.

**Figure S2.**
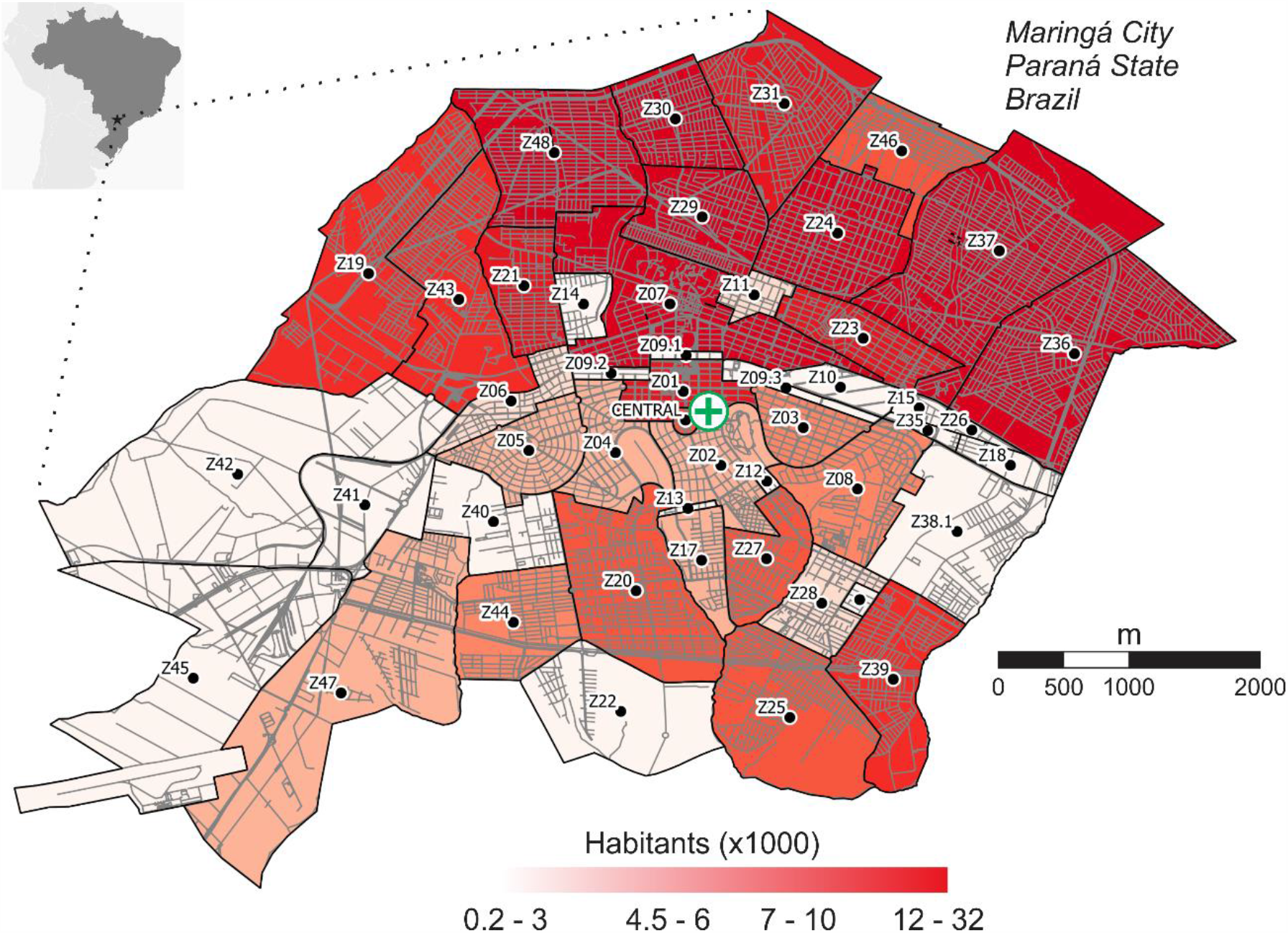
Location of the neighboring zones in the network city model - Maringá, PR, Brazil. Shades of red are scaled to the total number of citizens in each census zone, according to the 2010 national census. Roads and avenues are depicted by grey traces. The medical cross in the middle spots the neighborhood where the first COVID-19 case was empirically reported and inserted in each simulation.

**Figure S3.**
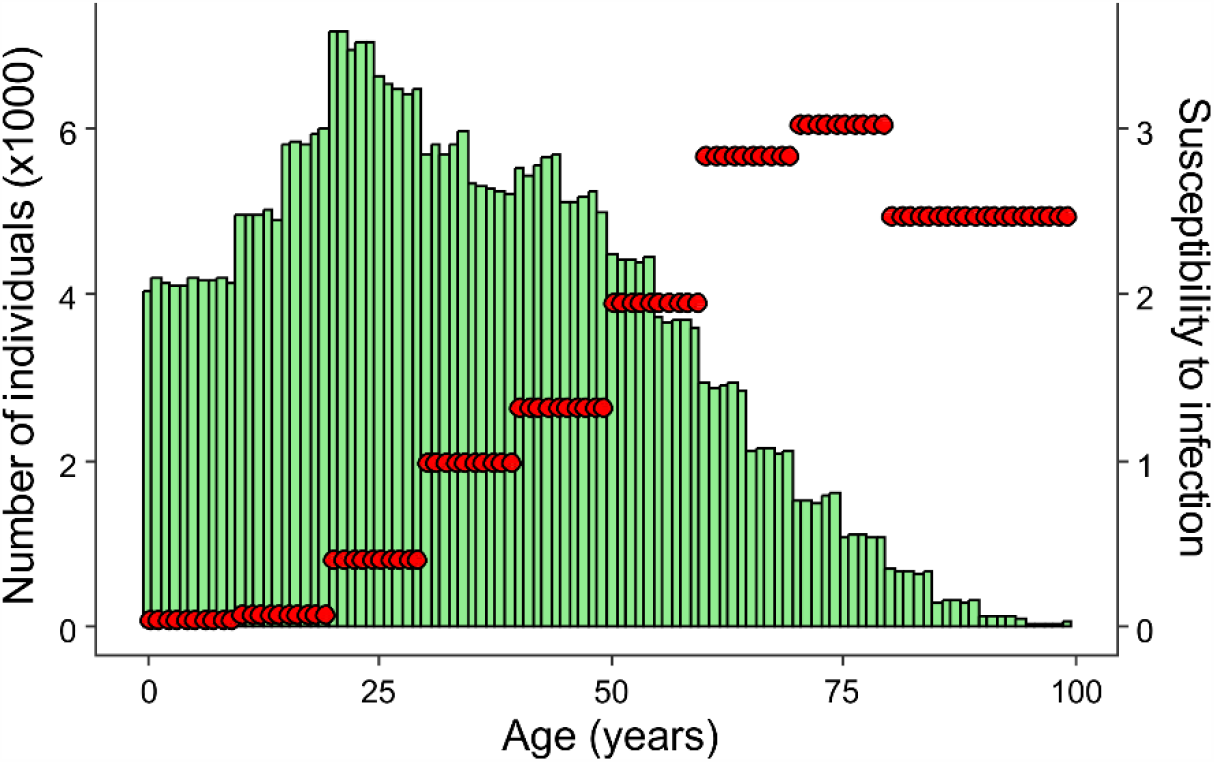
Age-specific numbers of individuals (green bars) and susceptibility to infection by SARS-CoV-2 (red circles). The age pyramid was obtained empirically using data on 357,077 citizens from the 2010 national census on our model city (Maringá, PR, Brazil). The age-dependent relative mean susceptibility to infection was obtained from Wu et al. (2020). The reference group to calculate values are people between 30 and 39 years, for which susceptibility is considered as 1.

**Figure S4.**
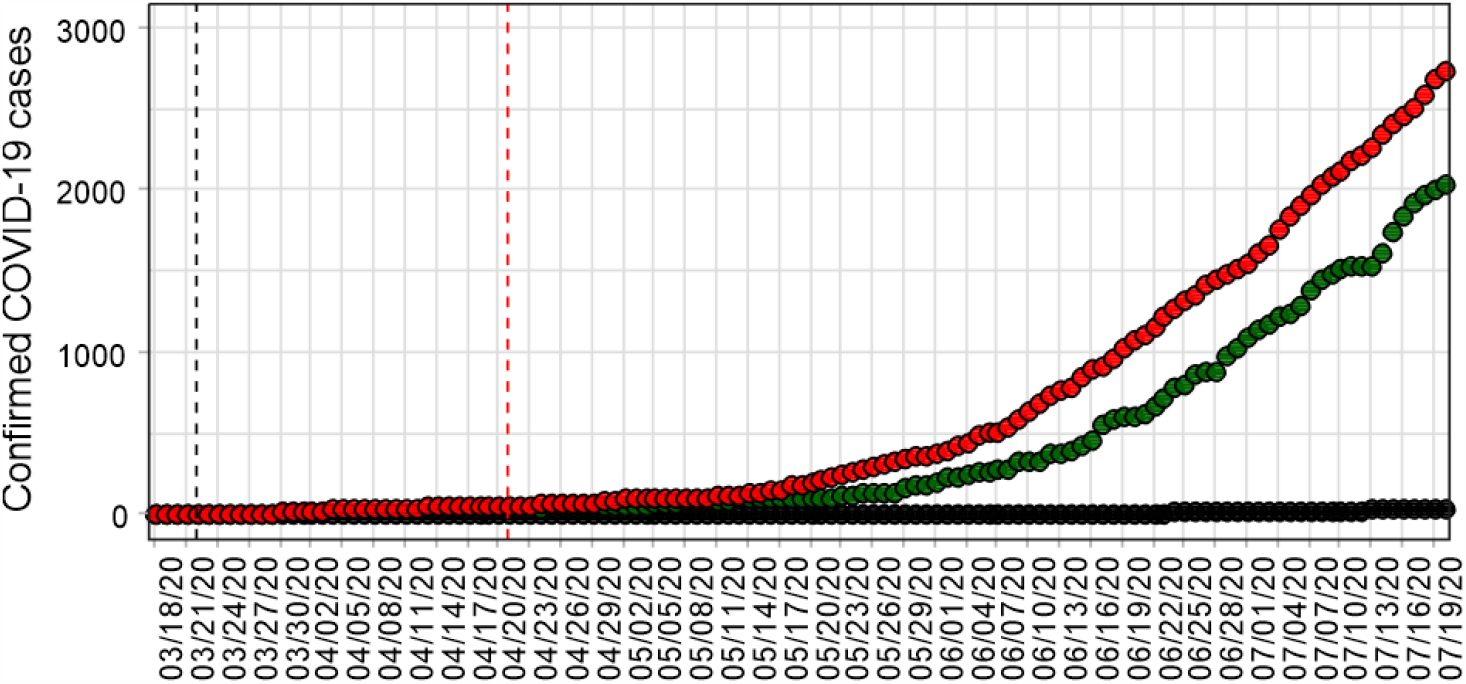
Time series of COVID-19 cases in the model city (Maringá, PR, Brazil). Red circles represent the number of confirmed cases, after clinical testing. Green circles depict the number of fully recovered cases, available back until April 14, 2020. Black circles represent the number of confirmed deaths. Vertical dashed lines are for the beginning of the social distancing protocol (black) and the start of the gradual recovery of working and social activities (red).

**Figure S5.**
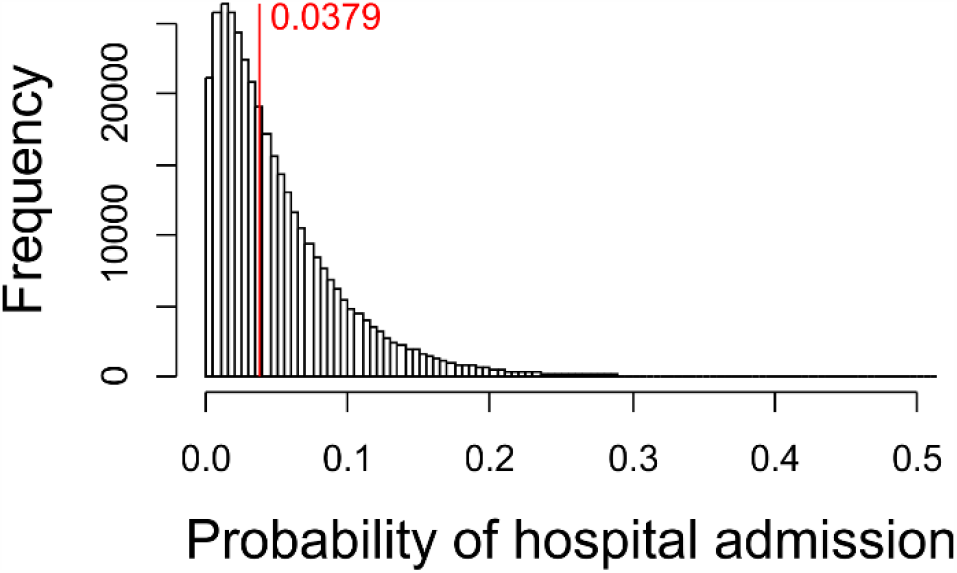
The estimated median probability of hospital admission. This probability combines the chance of an infected individual to develop severe/critical symptoms and the chance of requiring hospitalization such as internal assistance, intensive care, and mechanical ventilation (see *Empirical healthcare data*).

## Supplementary material

**Table S1.**
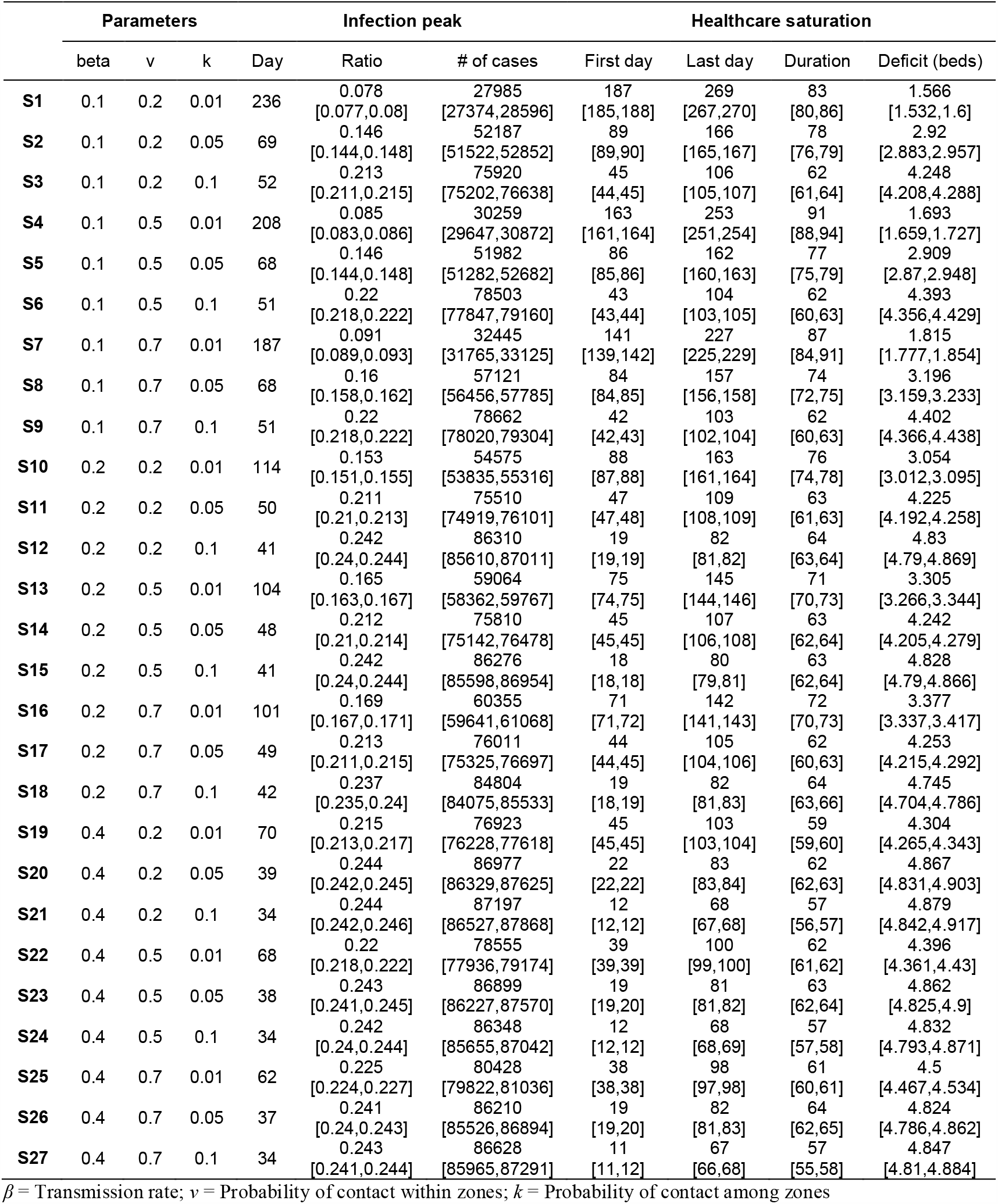
Model parameter and outputs with 95% confidence interval for the individual-based age-structured network model simulating the COVID-19 spread using Maringá, PR, Brasil as model city. Please refer to the full text for details.

## Notes

### Competing Interest Statement

The authors have declared no competing interest.

